# RESULTS OF A ONE YEAR STUDY OF THE STABILITY OF INTRAVENOUS NITROGLYCERIN IN 5% GLUTATHIONE FOR TREATING ADHF AND RIGHT HEART FAILURE POST LVAD IMPLANTATION

**DOI:** 10.1101/2025.03.24.25324579

**Authors:** Wayne Kaesemeyer

## Abstract

**BACKGROUND:** Superoxide causes diuretic resistance which limits the treatment of ADHF and right heart failure post LVAD implantation. Formulating IV GTN in 5% GSH is expected to benefit both these conditions with GTN targeting heart failure and GSH preventing diuretic resistance. The purpose of this study is to evaluate the stability of a new formulation of intravenous nitroglycerin (GTN) for use in treating ADHF and right heart failure post LVAD implantation.

**METHODS:** a new protocol for formulating IV GTN in 5% GSH that is buffered with 2% L-arginine was used to create two solutions of GTN in 5% GSH. Following this the pH of two solutions of .2 mg/cc GTN in 5% GSH prepared according to the new protocol was tested. The solutions were then stored at room temperature for one year and observed for evidence of GSH oxidation. After one year, the pH of the solutions was retested.

**RESULTS:** initial pH ranged 4.0 +/-.05 and was unchanged after one year. Both solutions remained clear without evidence of GSH oxidation.

**CONCLUSION:** this new formulation of IV GTN in 5% GSH is stable for use in treating ADHF and right heart failure post LVAD implantation for periods of up to one year. This new formulation has the potential to reduce the need for right ventricular assist devices following LVAD implantation and facilitate the explantation of LVADs thereby reducing the need for heart transplants.

**WHAT IS ALREADY KNOWN ON THIS TOPIC:** Diuretic resistance (DR) is known to be associated with increased heart failure mortality. DR may be increased by superoxide which is intrinsic to the pathophysiology of heart failure. GSH depletion from the use of intravenous nitroglycerin in the treatment of ADHF and RHF leads to superoxide production and DR, both of which may be GSH suppressible.

**WHAT THIS STUDY ADDS:** This study adds a new drug designed to prevent superoxide induced DR.

**HOW THIS STUDY MIGHT AFFECT RESEARCH, PRACTICE OR POLICY:** This study may lead to improved treatment of ADHF, prevention of RHF following LVAD implantation and facilitation of LVAD explantation for reducing the need for heart transplantation

## BACKGROUND

GTN was first discovered in 1847 by Sobrero (1). Tolerance seen in its treatment of patients was first described in 1888 (2). Over the ensuing years many hypotheses to explain and eliminate tolerance in the treatment of cardiovascular diseases have been advanced. And over the past 50 year there have been multiple studies showing the actions of GTN are GSH dependent and that GSH is involved in tolerance. However, it was only recently that superoxide from mitochondria in response to treatment with GTN was discovered (3). This was followed by the discovery GTN uncouples complex I resulting in superoxide formation which is suppressed by glutathione (4), along with tolerance.

GTN tolerance is a process that begins at mitochondrial complex I with its uncoupling by S-nitrosylation (4,5) from NO donated by and generated from GTN (6,7,8). The resulting superoxide formed depletes GSH by S-glutathiolation thereby leading to further superoxide which is GSH suppressible (4,9,10). This process ends at NADPH oxidase (figure)

In between mitochondrial complex I and NADPH oxidase there is uncoupling of aldehyde dehydrogenase II (ALDH II) and eNOS. eNOS uncoupling involves S-glutathiolation (11) and substrate depletion by arginase II (6,12), which is mediated by ROS and RNS formed following complex I uncoupling. This is followed by activation of protein PKC (6,13) which completes the process by activating NADPH oxidase with superoxide from a third and final site (figure).

Therefore, the site of formation of the first superoxide radical is mitochondrial complex I and NADPH oxidase is the site of the last superoxide radical’s formation. The net effect of superoxide from all 4 sites is formation of peroxynitrite (6,14) in place of NO from ALDH-2 and EDNO from eNOS. Tolerance with loss of vasodilator function is the result. This entire GSH dependent-reversible process is illustrated and summarized in the figure and discussed in complete detail in reference (6).

### GSH BASED INTRAVENOUS GTN FORMULATION REPLACING DEXTROSE BASED FORMULATIONS

Based on the foregoing discussion of studies supporting the mechanism of tolerance illustrated in figure 1 as rationale, a reformulation GTN in combination with GSH is proposed for intravenous use based on Table 1. Table 1 illustrates a new formulation of GTN for intravenous dosing that replaces 5% dextrose with 5% GSH. For maintaining pH in the range of 3-5, a requirement for preventing GTN degradation over time, 2 % L-arginine is used as a buffer. In addition to functioning as a buffer, L-arginine is anticipated to prevent eNOS uncoupling from system y+ transporter oxidative dysfunction leading to L-arginine depletion at the site of eNOS activity as a result of supplying L-arginine via passive diffusion as previously described (6,15,16). Therefore, L-arginine performs a dual-use function. This new IV formulation, similarly dosed as the current dextrose based GTN formulation, would replace that dextrose formulation in the treatment of acute decompensated heart failure and controlling right heart failure which complicates the use of left ventricular assist devices in the treatment of advanced heart failure, as previously discussed (6). Finally it should be noted that this new formulation’s calculated osmolality is 278 mOsms/L. After adjusting for the small amount of excipients from GTN, and the use of NAC as a preservative that prevents GSH oxidation, this new formulation is iso osmolar with serum which makes it suitable for long term continuous dosing leaving long term in-vitro stability as the question addressed by this study, with clinical effectiveness as a replacement for dextrose based formulations as questions to be addressed by studies that follow.

**Table 1.** Current reformulation of nitroglycerin in dextrose *5%* (A) and proposed reformulation of nitroglycerin in GSH (B)

| (A) Nitroglycerin in dextrose 5% |  |  |  |  |
| --- | --- | --- | --- | --- |
|  | Composition |  | Osmolarity (mOsmol/L)<br>(calc) | pH |
| | Nitroglycerin ($\mu\text{g}/\text{mL}$) | Dextrose hydrous, USP<br>(g/L) | | |
| 25 mg nitroglycerin in 5% dextrose injection | 100 | 50 | 428 | 4.0 (3.0–5.0) |
| 50 mg nitroglycerin in 5% dextrose injection | 200 | 50 | 440 | 4.0 (3.0–5.0) |
| 100 mg nitroglycerin in 5% dextrose injection | 400 | 50 | 465 | 4.0 (3.0–5.0) |
| (B) Proposed reformulation of nitroglycerin in GSH 5% |  |  |  |  |
|  |  |  | Osmolarity (mOsmol/L)<br>(calc) | pH |
| | Nitroglycerin ($\mu\text{g}/\text{mL}$) | GSH<sup>a</sup>, USP (g/L) | | |
| 25 mg nitroglycerin in 5% GSH injection | 100 | 50 | 428 | 4.0 (3.0–5.0) |
| 50 mg nitroglycerin in 5% GSH injection | 200 | 50 | 440 | 4.0 (3.0–5.0) |
| 100 mg nitroglycerin in 5% GSH injection | 400 | 50 | 465 | 4.0 (3.0–5.0) |

**Table 1** Current reformulation of nitroglycerin in dextrose 5% (A) and proposed reformulation of nitroglycerin in GSH (B)**(A) Nitroglycerin in dextrose 5%**
|  | Composition |  | Osmolarity (mOsmol/L)<br>(calc) | pH |
| --- | --- | --- | --- | --- |
| | Nitroglycerin ($\mu\text{g}/\text{mL}$) | Dextrose hydrous, USP (g/L) | | |
| 25 mg nitroglycerin in 5% dextrose injection | 100 | 50 | 428 | 4.0 (3.0–5.0) |
| 50 mg nitroglycerin in 5% dextrose injection | 200 | 50 | 440 | 4.0 (3.0–5.0) |
| 100 mg nitroglycerin in 5% dextrose injection | 400 | 50 | 465 | 4.0 (3.0–5.0) |

**(B) Proposed reformulation of nitroglycerin in GSH 5%**
| | Nitroglycerin ($\mu\text{g}/\text{mL}$) | GSH<sup>a</sup>, USP (g/L) | Osmolarity (mOsmol/L)<br>(calc) | pH |
| --- | --- | --- | --- | --- |
| 25 mg nitroglycerin in 5% GSH injection | 100 | 50 | 428 | 4.0 (3.0–5.0) |
| 50 mg nitroglycerin in 5% GSH injection | 200 | 50 | 440 | 4.0 (3.0–5.0) |
| 100 mg nitroglycerin in 5% GSH injection | 400 | 50 | 465 | 4.0 (3.0–5.0) |

**Figure 1.**
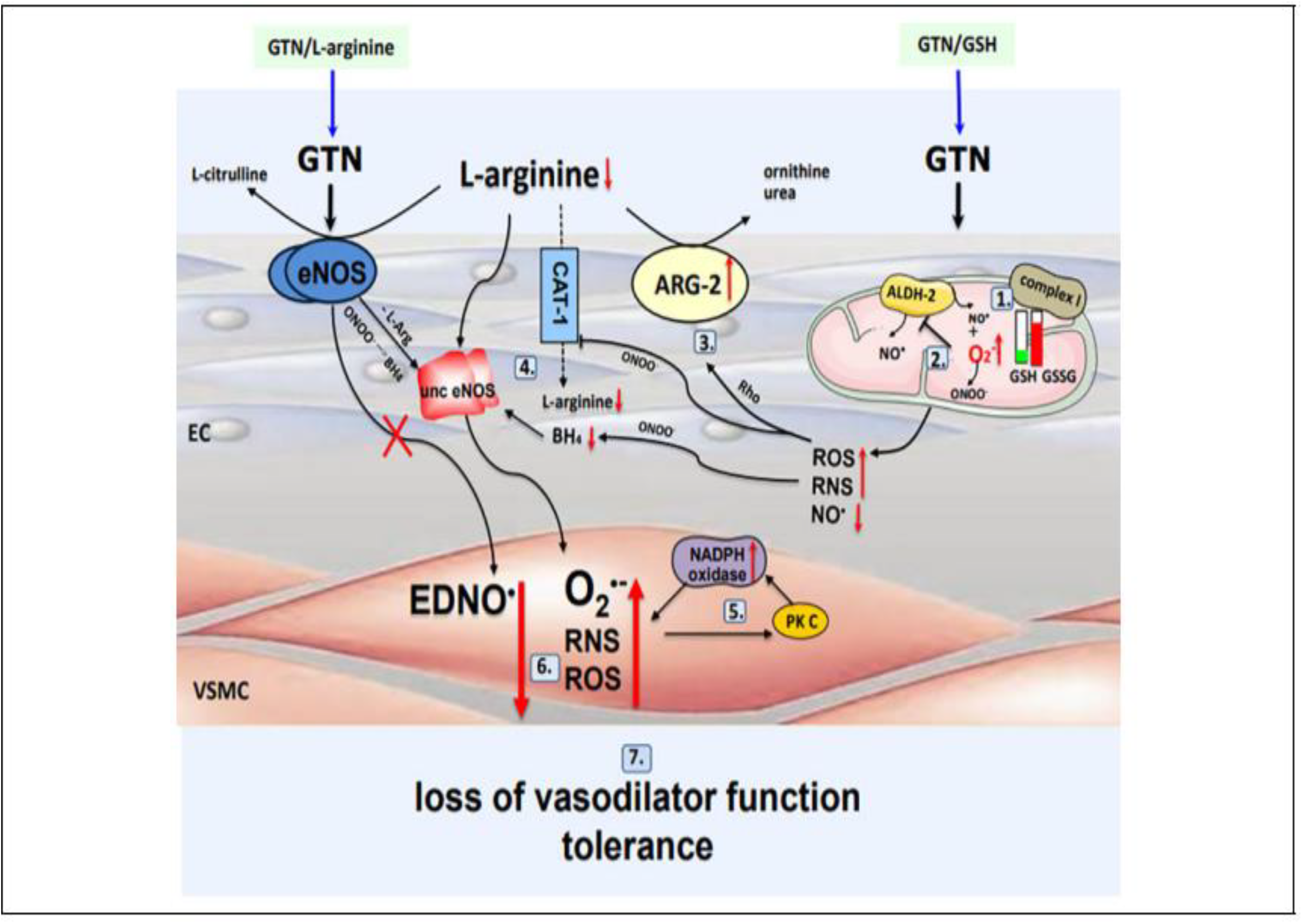
Cascading of superoxide mechanisms that mediate GTN tolerance. This 7 component process (I) begins at complex I. NO from GTN either from denitration or eNOS or both and leads to inhibition of complex I as a result of calcium dependent depletion of GSH and ROS and RNS generation. (2) ALDH-2 mediated bioconversion of GTN to NO is inactivated by ROS and RNS from step I, thereby leading to a loss of NO from bioconversion. (3) RNS from ALDH-2 upregulate arginase II via activation of RhoA/Rho kinase and reduce L-arginine availability to eNOS. (4) eNOS uncoupling from both. L-arginine and BH4 depletion further promotes superoxide production. (5) Superoxide from eNOS activates PK C and PK C subsequently activates NADPH oxidase. a fourth site of superoxide. (6) Superoxide from all 4 sites induces formation of peroxynitrite and related radicals in place of NO from ALDH-2 and EDNO from eNOS. (7) Tolerance is the end result of loss of vasodilator function. EC indicates endothelial cells; ARG-2. arginase-2; BH_4_, tetrahydrobiopterin; unc eNOS, uncoupled eNOS; GSH, glutathione; GSSG, glutathione di.sulfide; EDNO, endothelium-derived NO; PK C. protein kiinase C; ALDH-2, aldehyde dehydrogenase-2; VSMC, vascular smooth muscle cells; ROS, reactive oxygen species; RNS. reactive nitrogen species; CAT-I, cationic amino acid transporter I. 1.-7.-Summary items (see in text).

## METHODS

Beginning in August 2023 (8-21-23) two bottles, A and B, were formulated as described in Table 1. Both contained .2 mg/cc GTN, 5% GSH and 2% L-arginine. Bottle A differed from bottle B by the amount of NAC used to prevent oxidation of GSH to GSSG, .2% bottle A and .4 % bottle B. Following this both bottles were filtered using a 22-micron Millipore systems after which pH was measured by meter at a compounding pharmacy (Central Compounding, Durham, NC, USA) After this initial phase of the study both bottles were stored at room temperature for a year. After a year, on 8-28-24, the pH of both bottles was retested.

Materials : all agents used in formulations tested were USP pharmaceutical grade.

Nitroglycerin Injection, USP (5 mg/ml) was obtained from American Reagent, Inc., Shirley, NY 11967. A 22 micron Millipore system was used for sterile fill. pH was measured using a PH 90 EXTECH pH meter.

## RESULTS

8-21-23 - bottle A pH = 4.5 and bottle B pH = 4.0

8-28-24 – bottle A pH = 4.0 and bottle B pH = 3.88

Both 250 cc solutions remained clear and free of sediment as evidence of prevention of oxidation of GSH to GSSG by NAC (photo) and the formulation of .2 mg GTN in 5% GSH buffered with 2% L-arginine, using .2 - .4 %NAC as preservative was stable for use in heart failure studies for up to a year.

## DISCUSSION

### CONDITIONS THAT MAY BENEFIT FROM INTRAVENOUS GTN IN 5% GSH

Two difficult to treat conditions that may benefit from the above described intravenous GTN in 5 % GSH formulation are acute decompensated heart failure (ADHF) and right heart failure following LVAD placement. Both of these conditions have been treated with GTN with results considered negative due to the development of superoxide induced tolerance leading to diuretic resistance with continuous sustained GTN administration. Therefore, studies of the previously described I.V. GTN in 5% GSH based GTN formulation are proposed in the discussion that follows.

### BACKROUND FOR A STUDY OF THE IMPACT OF IV GTN in 5% GSH ON DIURETIC RESISTANCE IN PATIENTS WITH CONGESTIVE HEART FAILURE

The background for this is a recent study of cimlanod, a new nitroxl anion donor vasodilator being evaluated as a treatment for acute heart failure (17). The purpose of the study was to assess the effect of cimlanod on urine output and sodium excretion in response to diuretic treatment in stable patients with HFrEF with EF </= 45%. This study found that, patients with HF, congestion and a reduced LVEF, the infusion of cimlanod reduced urine volume and sodium excretion, both before and after the administration of furosemide, accompanied by plasma volume expansion and an increase in total body water, findings seen with other vasodilators, to include nesiritide, serelaxin, minoxidil, nitroprusside and IV GTN in 5% dextrose, studied in heart failure patients. As a result, it was concluded infusion of cimlanod attenuates a furosemide-induced diuresis in patients with heart failure and an LVEF <45%, which might have adverse consequences for those who are already congested, i.e. ADHF patients. Also, that development of interventions for acute heart failure should include research focused on assessing effects on water balance and potential interactions with diuretic agents.

### RATIONALE FOR A STUDY OF THE IMPACT OF IV GTN in 5% GSH ON DIURETIC RESISTANCE IN PATIENTS WITH ACUTE HEART FAILURE

Regarding the assessment of effects of vasodilators on water balance and interactions with diuretic agents, heart failure itself, unrelated to treatment, leads to increased renal superoxide levels (18). Therefore, those vasodilators previously mentioned that failed in previous clinical ADHF trials may have failed because of having no benefit on superoxide and diuretic responses or impaired responses leading to diuretic resistance (cimlanod).

As discussed previously (6), cimlanod is a nitroxyl anion donor and and nitroxyl (HNO) reacts with molecular oxygen and forms peroxynitrite at physiological pH (19). Therefore, one can view cimlanod as a peroxynitrite prodrug. Furthermore, peroxynitrite is a highly reactive radical which can uncouple eNOS by oxidizing BH4 like superoxide from which it can be formed. All of this as discussed below explains cimlanod’s effect to blunt Na+ and water excretion as in the above discussed study (17).

Mitochondrial superoxide has been found to reduce urine flow and sodium excretion in a rat kidney study (20). And this was prevented using a superoxide dismutase mimetic. In so far as GSH similarly suppresses mitochondrial superoxide from GTN induced mitochondrial complex I uncoupling, it is proposed that IV GTN in 5% GSH be used in place of cimlanod in a repeat of the cimlanod diuretic study (17).

This would be another short 8 hour infusion study in stable HF patients with reduced ejection fraction receiving loop diuretics. Patients would be randomized to infusion with IV GTN in 5% GSH vs matching infusion of IV GTN in 5% dextrose on separate days. Each infusion would be dosed to lower systolic BP by 20 mm Hg. during a short run-in period. At 4 hours following completion of the run-in period patients would receive 40 mg furosemide by IV bolus infusion. The primary endpoint would be urine volume at the end of 8 hours following completion of the run-in period, along with total Na+ excretion, fractional Na+ excretion, estimated glomerular filtration rate, systolic BP and hemoglobin level.

The purpose of this study would be to evaluate the effect of IV GTN in 5% GSH on sodium excretion and diuretic responses in stable diuretic treated heart failure patients with reduced ejection fraction. Reducing diuretic resistance in diuretic treated heart failure patients would be a significant improvement.

As for a reason to anticipate a positive response to IV GTN in 5% GSH, this is based on the positive effect seen with the SGLT2 inhibitors in acute heart failure. Similarly to GSH, dapagliflozin, a SGLT2 inhibitor, has been found to reduce oxidative stress in human proximal tubular cells (21). This may explain the results of empagliflozin, another SGLT inhibitor, on the response to diuretics and urine output seen in the EMPAG-HF trial (22), all of which suggests a rationale for combining these two agents in the treatment of ADHF. That is, SGLT2 inhibitor plus IV GTN in 5% GSH in combination initially, reserving IV diuretics for congestion unrelieved by GTN.

A second reason to anticipate a positive response in the above study is superoxide intrinsic to heart failure leads to insulin resistance (23). And, insulin promotes Na+ retention by the distal tubule thereby decreasing distal tubule Na+ excretion, a second site of DR (24,25). Therefore, by suppressing superoxide GSH reverses both IR and DR from decreased distal tubule Na+ excretion, in addition to reversing DR involving the proximal tubule described above, i.e. dual target, proximal and distal tubule, DR reversal by GSH.

Consistent with all of this are recent studies of patients treated for MI with PCI. In one study 25 g of GSH administered over 10 minutes on three consecutive days had a favorable effect on contrast associated AKI (26). And a benefit on LV remodeling post MI, using the same three day protocol, was seen in another study (27).

Lastly, as a possible future study going back to back to dapagliflozin discussed above (21), it should be noted that its solubility of .5 mg/ml aqueous buffer with pH 7.2, or less, may enable adding it to the formulation of GTN in 5% GSH that is described in table 1. If this approach is taken each 250 cc bottle of IV GTN in 5% may only need 10 mg of dapagliflozin, the current daily dose of that drug. This would avoid the need for the use of two separate drugs as described above for targeting DR in the treatment of ADHF.

### TREATING RIGHT HEART FAILURE FOLLOWING LEFT VENTRICULAR ASSIST DEVICE PLACEMENT WITH IV GTN in 5% GSH

Similarly to CHF from left heart failure, RHF may be another disease with mitochondrial superoxide involved in its pathogenesis. A recent RHF pulmonary artery banding study done in mice revealed the transition of maladaptive RV remodeling into RHF involved mitochondrial superoxide (28).

In patients requiring LVADs for treatment of advanced HF, there is the additional problem of RHF from pulmonary hypertension. RHF was seen in upwards of 20% of patients in the MOMENTUM-3 study. The use of an RVAD was required in 4% (29).

Multiple approaches for treating pulmonary hypertension leading to RHF seen with LVADs have been tried with limited benefit. This includes inhaled nitric oxide, IV GTN in dextrose 5%, nitroprusside, inotropes, sildenafil and endothelin antagonists. The vasodilator most frequently used has been nitroprusside.

Recently higher preoperative diuretic dose has been found to be associated with risk of early RHF following LVAD surgery (30). Therefore, IV GTN in 5% GSH would seem to be the next logical vasodilator that could be considered on the basis that superoxide may have limited previous attempts of diuretics to control RHF or promoted RV remodeling requiring RVAD placement. That is, GTN to control RHF and GSH to suppress superoxide that causes diuretic resistance

As for using IV GTN in 5% GSH to reduce DR and RHF post LVAD implantation seen in the study by Huang (30) this paper proposes minor changes in that study’s protocol. That study divided diuretic responsiveness into tertiles in terms of dose requirements in over a 72-hour period prior to LVAD implantation and concluded that the risk of early RHF following LVAD surgery is directly proportional to the preoperative diuretic dose. Therefore, it is proposed that at the beginning of the 72-hour pre -LVAD implantation period, prior to beginning of the diuretic efficiency assessments, an infusion of IV GTN in 5% GSH, 0.2 mg/cc GTN, be started at 10 mcg/min followed by a gradual up titration in the GTN dose to optimize right atrial (RA) pressure, pulmonary artery systolic pressure, pulmonary artery diastolic pressure, pulmonary capillary wedge pressure (PCWP), and cardiac index, parameters for RHF control, at which point the GTN dose would be fixed for the remainder of the 72 hours. At that point the remainder of the previous protocol would be followed for studying DE with results grouped again into tertiles with need for RVADs and mortality reassessed with reconstruction of figures 2 and 3 plus Tables 2 and 3.

For those patients who have responded favorably with extended use of LVADs there is a need to improve the process for explantation to reduce the need for heart transplantation. Currently up to five oral drugs are required to maintain patients hemodynamically stable during the weening process which is successful in about 50 % of attempts after 18 months (31). Therefore, this paper proposes studying the use of IV GTN in 5% GSH to reduce the need for multiple oral agents controlling RHF and to reduce weening time.

## CONCLUSIONS

1] 0.2 mg/cc GTN in 5% GSH buffered with 2 % L-arginine has been found to be stable for one year.

2] Benefits of this new formulation of IV GTN on reducing diuretic resistance seen in the treatment of acute heart failure, right heart failure following LVAD placement and facilitation of explantation of LVADs to reduce the need for heart transplants remain to be determined in future studies.

## Data Availability

available on request

## ABREVIATIONS

ADHF: acute decompensated heart failure
AHF: acute heart failure
BH4: tetrahydrobiopterin
DR: diuretic resistance
eNOS: endothelial nitric oxide synthase
GSH: glutathione
GTN: nitroglycerin, glycerol trinitrate
IV GTN/GSH: intravenous nitroglycerin in glutathione 5%
IR: insulin resistance
LVAD: left ventricular assist device
NAC: n-acetylcysteine
NO: nitric oxide OONO-peroxynitrite
PKC: protein kinase C
RHF: right heart failure
ROS: reactive oxygen species
RNS: reactive nitrogen species
RVAD: right ventricular assist device
SGLT2: sodium glucose transporter 2

## DECLARATIONS

## Funding

(information that explains whether and by whom the research was supported) – N/A. There was no funding for this manuscript

## Conflicts of interest/Competing interests

(include appropriate disclosures) – N/A The author has no conflicts to declare.

## Availability of data and material

(data transparency) – available on request

## Code availability

(software application or custom code) – Microsoft word

## Authors’ contributions

the author of this paper researched, organized, wrote and proofread the paper as its sole author.

## Ethics approval

(include appropriate approvals or waivers) – N/A The manuscript did not involved studies of animal or human subjects

## Consent to participate

(include appropriate statements) – N/A

The manuscript did not require consent to participate as it did not involve studies of animal or human subjects

## Consent for publication

(include appropriate statements) – the author of the paper consents in its publication.

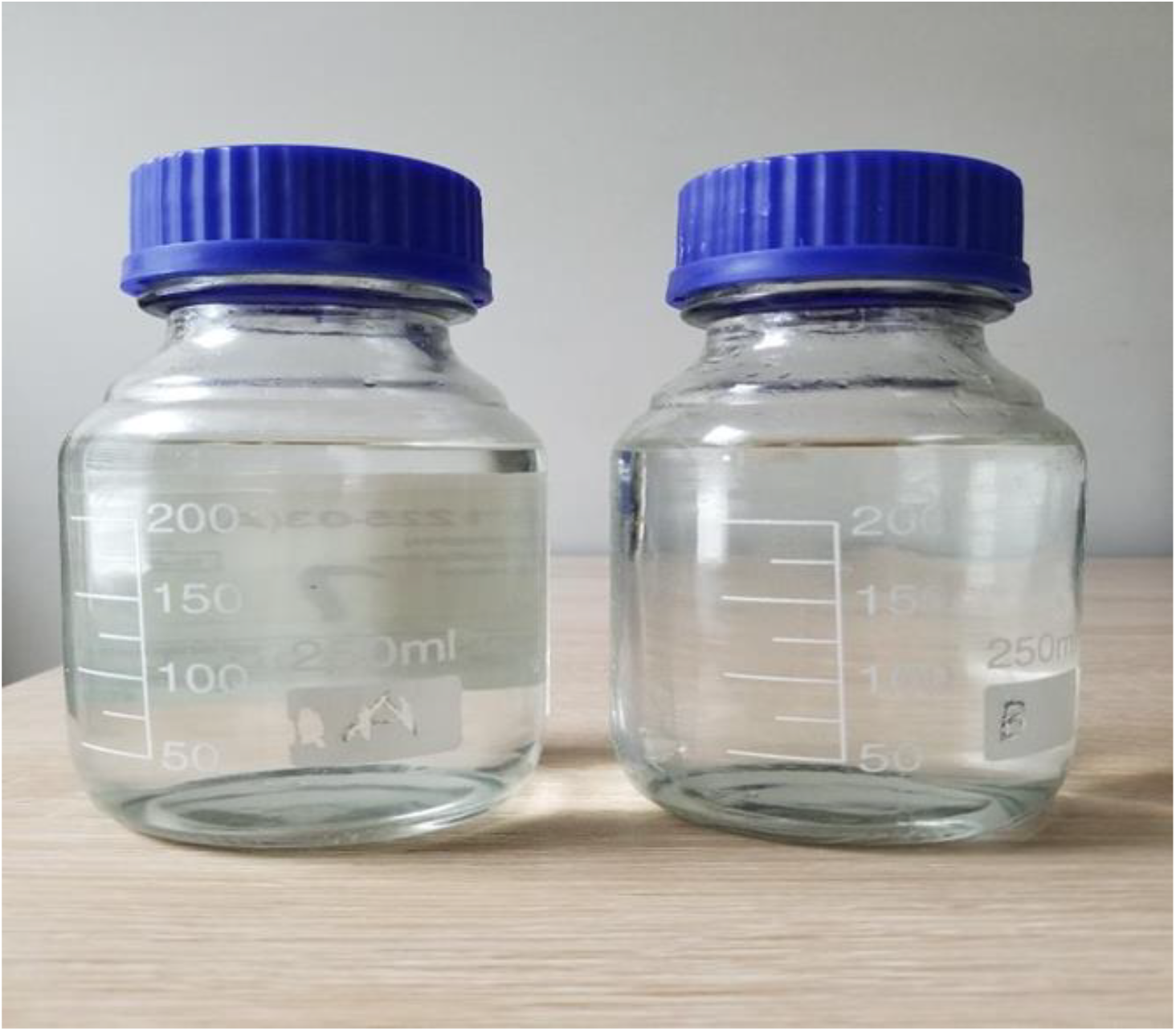

Legend for photo. Bottles A (left) and B (right) on 12-20-24. Both formulated on 8-21-23, stored at room temperature, and re-tested on 8-28-24. See text for details.

## Notes

Sources of support: none

Conflicts: none to declare

### Competing Interest Statement

The authors have declared no competing interest.

### Clinical Trial

n/a

### Funding Statement

Funding for this study provided solely by author

### Author Declarations

IRB approval not applicable - animal and human subjects were not involved

